# Uncertainty and Value of Information Analysis in the Integrated Transport and Health Impact Modelling Tool for Global Cities (ITHIM-Global)

**DOI:** 10.1101/2025.08.15.25333754

**Authors:** Anna Schroeder, Lambed Tatah, Ali Abbas, Rob Johnson, Haneen Khreis, Christopher Jackson, James Woodcock

## Abstract

Health impact assessment models are a key tool for stakeholders and policymakers to understand the effect of transport on public health. The Integrated Transport and Health Impact Model for Global Cities (ITHIM-Global) is an open-source model developed specifically for low- and middle-income countries to assess the impact of changes in transport on population health at a city level. It models impacts through three different pathways: air pollution, physical activity and road traffic fatalities. As there is uncertainty in input parameters, ITHIM-Global can be set up to use a Monte Carlo simulation, sampling from pre-defined probability distributions describing inputs to create credible intervals for the various model outputs. A Value of Information analysis is used to determine the effect of these various input parameters on the uncertainty of the outputs. Using Bogotá as an example, this article explains how this uncertainty analysis works in ITHIM-Global. We demonstrate the effect of uncertainty on estimates of years of life lost at a city level using three hypothetical scenarios resulting in 5% increases in cycling, public transport and car journeys, respectively. The use of a Value of Information analysis and the impact of the various input parameters on the uncertainty in the results is also demonstrated. We show that an increase in cycling or public transport always has a positive effect on population health, independently of the uncertainty in our input parameters, whereas an increase in car journeys always has a negative impact. We also show that having perfect knowledge of some of the input parameters individually could reduce the standard deviation of the credible intervals of our results by up to 7%.

## 1. Introduction

Through many interacting pathways, such as physical activity and air pollution, and traffic injuries, transportation has a large impact both positively and negatively on the health of a population (1–3). Health impact models (quantitative health impact assessment) provide a key tool for stakeholders and policymakers to understand these impacts and how potential changes in transport planning and policy can impact population health (4).

According to a recent review (5), the two most commonly used health impact models between 2014 and 2020 were the Integrated Transport and Health Model (ITHIM) (6) and the WHO’s Health Economic Assessment of Transport (HEAT) tool for walking and cycling (7).

ITHIM estimates the impact of transport on the population health of a city via the pathways of air pollution, physical activity and road-traffic fatalities using location-specific inputs. There are different versions of ITHIM (6,8–12). ITHIM-Global was developed to model the health impacts of transport in low- and middle-income countries where data tend to be more scarce (13), leading to significant uncertainty in the input data.

The consequences of uncertainty in the input parameters used in health impact models are often explored by a one-way sensitivity analysis, i.e. examining how a model output changes with different potential input parameter values (12,14). While this can give an indication of the model’s sensitivity to its inputs, it relies on arbitrary choices and can be difficult to interpret if there are correlations between parameters or nonlinear relationships between parameters and outcomes. A more advanced approach is to use probabilistic uncertainty analysis, which is a way to deal with uncertainty in multiple input parameters at the same time (15). This methodology requires defining probability distributions for the relevant input parameters that represent the strength of evidence for each one and then using a Monte Carlo simulation: running the model many times, each time sampling the input parameters from their respective distributions. This produces distributions that represent plausible values for the model outputs.

Following a probabilistic uncertainty analysis, Value of Information Analysis (VoI) can be used to determine the influence of the uncertainty in the various input parameters on the uncertainty in the output values (16). VoI methods determine the expected benefits of further knowledge of various kinds. It gives us the means to determine by how much we might expect to reduce our uncertainty in the output values if we knew the true value of an input parameter.

In this paper, we introduce uncertainty and VoI analysis within the ITHIM-Global model, using the city of Bogotá (Colombia) as an example. We consider the health outcomes (Years of Life Lost (YLLs)) of three hypothetical scenarios assuming a 5% mode shift towards public transport, cars (private vehicles) or cycling. We demonstrate how we can build input parameter distributions, and how, by running the model many times, each time sampling from these distributions, we can calculate credible intervals for our output values. We also determine the impact of each of these parameters on the output values via a VoI analysis. This is done by calculating the *Expected Value of Partial Perfect Information* for the model parameters, a quantity that captures the extent to which we could reduce the uncertainty in our results were we to know the true value of an input parameter.

This approach enables us to represent and evaluate uncertainty in data in low- and middle-income countries as ITHIM-Global relies on many input sources, which may not be that reliable. Having a robust model to understand the impact of transport on public health that explicitly incorporates uncertainties in inputs helps to reduce health inequities between the Global North and South.

## 2. Description of ITHIM-Global

ITHIM-Global is an open-source, modular model designed to estimate the health and environmental impacts of shifts in urban passenger transport modes, particularly in low- and middle-income countries, where data availability and policy relevance are especially critical, on population health. ITHIM-Global simulates changes in all-cause and cause-specific mortality and years of life lost (YLL), alongside carbon dioxide emissions, using a comprehensive framework that integrates three primary exposure pathways: physical activity, air pollution, and road traffic injuries. The model employs a quasi-microsimulation approach, assigning individualised exposures by age, sex, and activity level to generate nuanced, population-specific estimates. This individual-level exposure assignment enables the model to utilise the most up-to-date exposure- and dose-response functions, which are particularly important for capturing non-linear relationships such as those between physical activity and health.

Central to the ITHIM-Global model is its use of detailed, city-level input data. These include household travel surveys, physical activity surveys, air pollution and vehicle emissions data, road traffic injury records, and demographic and mortality statistics. Data harmonisation and pre-processing routines are employed to address inconsistencies, ensure comparability across settings, and accommodate typical data gaps in low- and middle-income cities, such as incomplete travel stage information or missing trip characteristics. When local data are unavailable or incomplete, ITHIM-Global allows the integration of global parameters and imputation methods, providing a pragmatic yet robust approach for diverse urban contexts.

Scenarios are implemented by specifying counterfactual changes in travel mode shares while holding the total number of trips constant. To demonstrate model functionality, the ITHIM-Global team applied three hypothetical scenarios in Bogotá, each increasing the share of trips by five percentage points for cars, buses, or cycling, respectively. Trips are stochastically reassigned to the target modes based on trip distance distributions observed in the underlying travel survey to enhance realism. The effects of these scenarios are assessed at both the individual and population levels, capturing the resultant shifts in physical activity, exposure to air pollution, and risk of road traffic fatalities and attributable health impacts.

The model creates a large set of outputs. Health impacts are expressed as changes in all-cause and cause-specific mortality and YLLs, with results disaggregated by pathway and disease grouping. Outcomes are organised into three levels: overall (all-cause), disease group (such as cardiovascular disease or cancer), and specific diseases (e.g., type 2 diabetes, stroke). All three levels also include road traffic fatalities (see S8). Environmental impacts are captured through estimates of transport-related CO₂ emissions. The modular architecture of ITHIM-Global supports extensive sensitivity and uncertainty analyses.

Recent innovations distinguish ITHIM-Global from previous iterations, including more granular exposure estimation, harmonised dose-response relationships predicting relative risks for various disease outcomes for each person in our model population, enhanced modelling of travel patterns, and robust open-access documentation and code. A large number of input parameters, which can be set up as probability distributions, make the tool particularly well-suited for VoI analyses, as it allows explicit parameterisation of key uncertainties, transparent sensitivity analyses, and systematic quantification of the influence of input parameter uncertainty on model outputs. By integrating flexible data requirements and modular scenario analysis, ITHIM-Global provides an accessible yet sophisticated platform for quantifying alternative urban transport policies’ potential benefits and uncertainties. For a more detailed description of ITHIM-Global, please see (13). The open-source model code for ITHIM-Global, called ITHIM-R, can be found on Github (17).

## 3. Dealing with uncertainty in the input parameters

While the model is by nature an approximation of a complex real-world process, it allows a wide range of uncertainties to be represented through its input parameters. In this section we explain how probabilistic uncertainty quantification is used to determine the uncertainties around model outputs.

### 3.1 Monte Carlo simulation and 95% credible intervals

In transport health impact modelling, parameters are commonly uncertain for a variety of reasons. Data might not be available for the city of interest, or only available for a small and/or selected sample of individuals (data uncertainty). There is also data variability between individuals. We generally do not include variability between individuals in the model apart from the variability found in the travel, physical activity and road traffic fatality surveys.

To quantify the impact of the uncertainties, many models use a sensitivity analysis (12,14) to calculate the range of expected outcomes when a single input parameter varies between two extremes, usually the upper and lower ends of an (estimated) 95% credible interval. While this gives evidence about the impact of each parameter separately, the choice of extreme values is arbitrary and has the potential to conceal nonlinear changes, and the joint impact of all input uncertainties is not represented.

In ITHIM-Global we use a different approach that allows us to calculate 95% credible intervals for our various outputs by including uncertainty in many of our parameters at the same time. We define probability distributions for most of the input parameters, such that these distributions represent our knowledge and judgement about the relative plausibility of the different values each parameter could take. By running the model many times, each time using different samples from those distributions, i.e. by performing a Monte Carlo simulation, we can calculate a range of potential output values. This method allows us to determine the consequences of the uncertainties in all the input parameters at the same time and quantifies the overall strength of evidence for the model outputs.

### 3.2 Introduction to Value of Information Analysis

After setting up input parameter distributions and performing a Monte Carlo simulation, analysts can perform a Value of Information (VoI) analysis to assess how the input distributions impact output uncertainty. This section gives a brief introduction to VoI analysis, and in particular to the concepts used within ITHIM-Global. For a more detailed and general introduction, please see (16).

VoI analysis enables us to determine the extent to which the uncertainty in each input parameter contributes to the uncertainty of the output values. It quantifies the expected reduction in uncertainty, i.e. the reduction in variance or standard deviation, of an output value if we were to obtain better (more precise) information about the different input parameters.

Let *θ* denote the set of input parameters and let f() be the deterministic function representing the model, i.e. Y = f(θ). The expected model output, with respect to uncertainty about the model inputs, is *E*_θ_(f(θ)). ITHIM-Global is run many times, each time using new samples from the input parameter distributions of *θ*, i.e. a Monte Carlo simulation is used. The expected outcome *E*_θ_(f(θ)) is estimated as the mean of the outcomes of these runs, and the uncertainty about the output can be quantified by computing a 95% credible interval from the sample of outputs. Note that *E*_θ_(f(θ)) might be different from the f(θ) we would calculate if we knew the true θ. We want to know which of the parameters comprising θ have the biggest impact on the uncertainty of the output. This allows us to prioritise future research on the parameters that have the largest impact.

The *Expected Value of Partial Perfect Information* (EVPPI) is defined in this context as the expected reduction in the variance of the model output if we were to learn the true value of an uncertain parameter, such as the “chronic disease scalar”. The chronic disease scalar represents uncertainty we have about the relevance and accuracy of Global Burden of Disease data (18) which we use for our baseline mortality statistics. All estimated deaths and YLLs for each disease outcome of interest are multiplied by this scalar during an ITHIM-R run. Let the chronic disease scalar be θ_1_in θ. Then

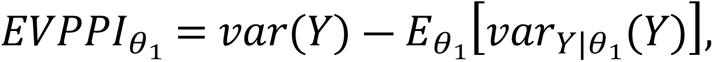

Where 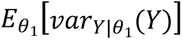 is the expectation of the variance of the model outputs if we knew θ_1_, the chronic disease scalar, exactly. Note that we take the expectation of 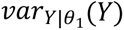 rather than 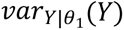 itself because we do not know θ_1_ for certain.

In practice, the 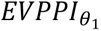 is calculated as follows. Let *R* be the number of times ITHIM-Global is run in the Monte Carlo procedure, 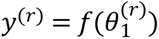 for each *r =* 1, …., *R.* Let *y^(r)^* be the total of all health outcomes, i.e. the number of YLLs gained, in the public transport (PT) scenario, i.e. when 5% of non-PT trips are converted to PT trips. Then *y^(r)^* can be approximated with a regression model

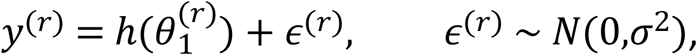

where 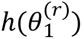 is the fitted function and ϵ^(*r*)^ are the residuals as can be seen in Fig 1.

**Fig 1.**
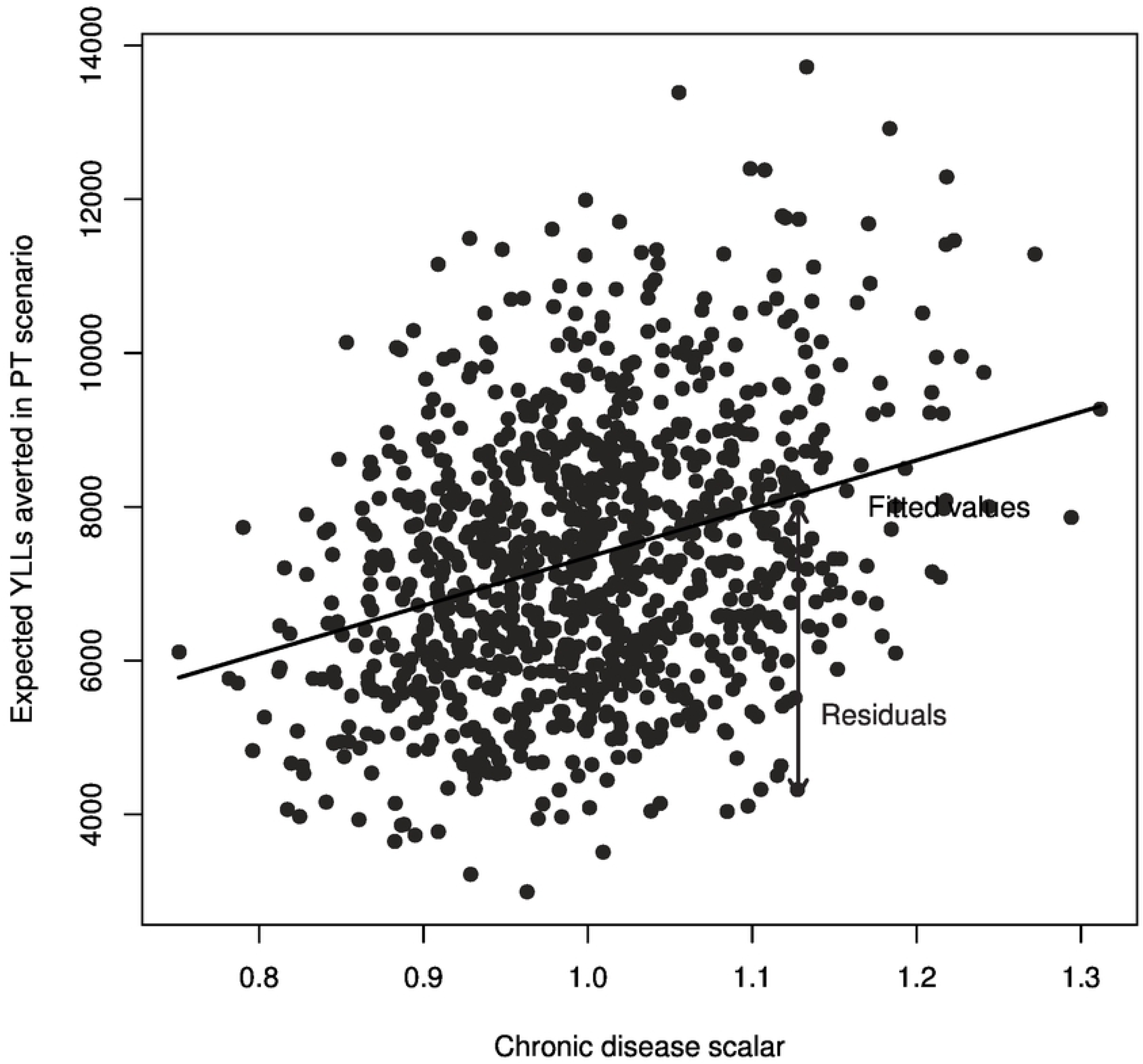
Model outputs versus the input values of the chronic disease scalar.

The value of the fitted function 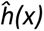 for an input value *x* is *E*(Y| θ_1_ = *x*), the expected outcome if the uncertain parameter θ_1,_ were known to equal *x*. Note that we use 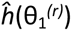 to represent our estimate of the true function *h*(θ_1_*^(r)^*). The residuals 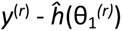 represent the variability in the outcome that is not explained by θ_1_, our chronic disease scalar.

The expected value of partial perfect information that can be gained by knowing θ_1_ explicitly is given as 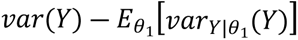. This can be estimated by subtracting the residual variance from the variance of *y*^(*r*)^, i.e.:

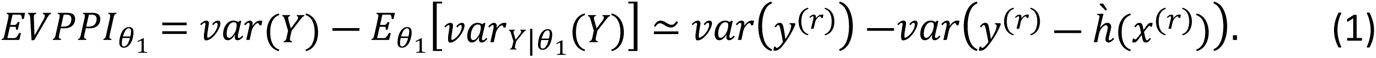

In some cases, the input parameters do not have independent effects on the output uncertainty, e.g. the percentage contributions of the various modes to the total PM_2.5_ emissions. In these instances, we want to know the EVPPI corresponding to learning multiple input parameters concurrently. Here, we use a nonparametric regression with more than one predictor, i.e.

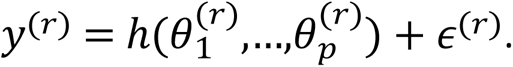

*h*() is a flexible, non-linear function; for example, a generalized additive model based on splines (16). By calculating the EVPPI for different outputs and parameters, we can identify places in the model where better information about a specific parameter might lead to improvements in the precision of the model outputs. For parameters with low EVPPI, we can conclude that further research is not worthwhile. For those with high EVPPI, further research *may* be worthwhile; given that research will not produce perfect information, the EVPPI is an upper bound for the potential value of this research. The expected benefits of a specific research study could be determined with the expected value of *sample* information (EVSI), though this is not considered in this paper (16).

### 3.3 Value of Information Analysis in ITHIM-Global

The ITHIM-Global model allows the user to create distributions for a wide range of input parameters. The model is set up to sample from these input parameter distributions anew each time the model is run. By running the model many times, i.e. by using a Monte Carlo simulation, we create a distribution of output values from which we derive credible intervals for our reported results as well as the EVPPI of the input parameters.

EVPPI values for input parameters may vary between the different combinations of outputs and scenarios. In this work we consider all possible combinations for years of life lost outcomes. This comprehensive approach allows us to determine which input parameters have the most impact across all outcomes of interest, and thus identify which parameters present the most potential as candidates for future research, with the aim to reduce our uncertainty about them in order to ultimately reduce output uncertainty.

The result of the VoI analysis is given as the proportion of output standard deviation that we could eliminate by learning each parameter. We first find the proportion of the variance (using Eqn (1)):

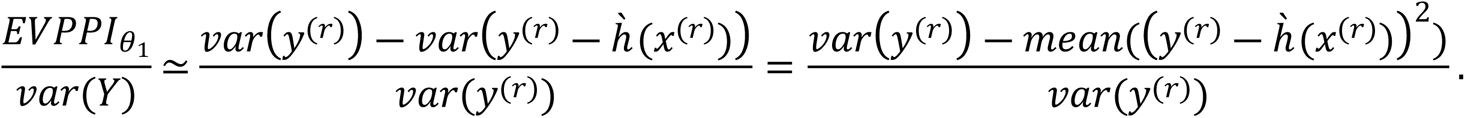

This holds as 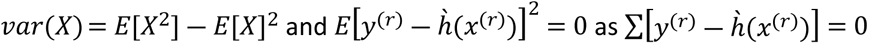 by the nature of regression functions. We find standard deviations to be a more intuitive representation of uncertainty, and therefore, for the purposes of presentation, we calculate the corresponding reduction in standard deviation by taking the square root, 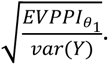

In ITHIM-Global, most input parameters are not correlated with other input parameters. We assume them to have independent effects on the model outputs and therefore run the VoI analysis individually for those input parameters. For some input parameters, we do not set up confidence intervals directly but rather compute probability distributions during a model run (e.g., the proportion of people without non-travel physical activity – see S6.3 for more details). In those cases, the VoI analysis is performed on the sampled values calculated within the model.

The VoI R package developed by (19) is used to calculate the EVPPI for each input parameter for which a probability distribution is defined. For single-parameter analyses, we use a nonlinear regression method based on nonparametric regression as described by (20). For interdependent input parameters, a generalised model based on linear splines (19) is applied using the ‘earth’ package (21). We present results generated using 5000 samples (see S11 for an analysis of model output and VoI estimates’ sensitivity to sample size).

S3 – S7 explain how probability distributions are set up for the various input parameters in this application. S9 explains how to conduct Monte Carlo simulation and VoI analysis with ITHIM-R.

## 4. Input parameter uncertainty quantification

There are different ways to choose the input parameter distributions for ITHIM-Global. In S3-S7 we demonstrate how we decided on the parametrisation of distributions in detail for the case of Bogotá. Here, we identify some general principles which can be used when defining such distributions:

- start with statistical sampling uncertainty / published standard error or confidence intervals where available and applicable.
- for most parameters, uncertainties arise from a lack of data that correspond directly to the modelled context (e.g. city, exposure definition). As such, no appropriate published standard error or confidence interval is generally available to quantify uncertainty – instead, judgements are required.
- sometimes a range of values for different contexts (e.g. locations, or ways of measuring exposure) are available. Credible intervals for the modelled context are then chosen to span these different estimates.
- without precise knowledge of the extent of uncertainty, we aim to be conservative and err on the side of wider credible intervals. Then, if the parameter is deemed to be not influential, the model’s conclusions are strengthened and there is little need to further refine the range.
- bias parameters are used to adjust data to represent the modelled context if we have some doubt in the data. Given lack of knowledge of the size and direction of this bias, these parameters are chosen with a credible interval centred around no bias (e.g. representing +-5% bias)

Table 1 gives a summary of the input parameters for which uncertainty is quantified, and their associated credible intervals. For an explanation of how these credible intervals were derived, and how probability distributions were defined from them, see S3-S7. For all remaining input parameters please see S1.

**Table 1.**
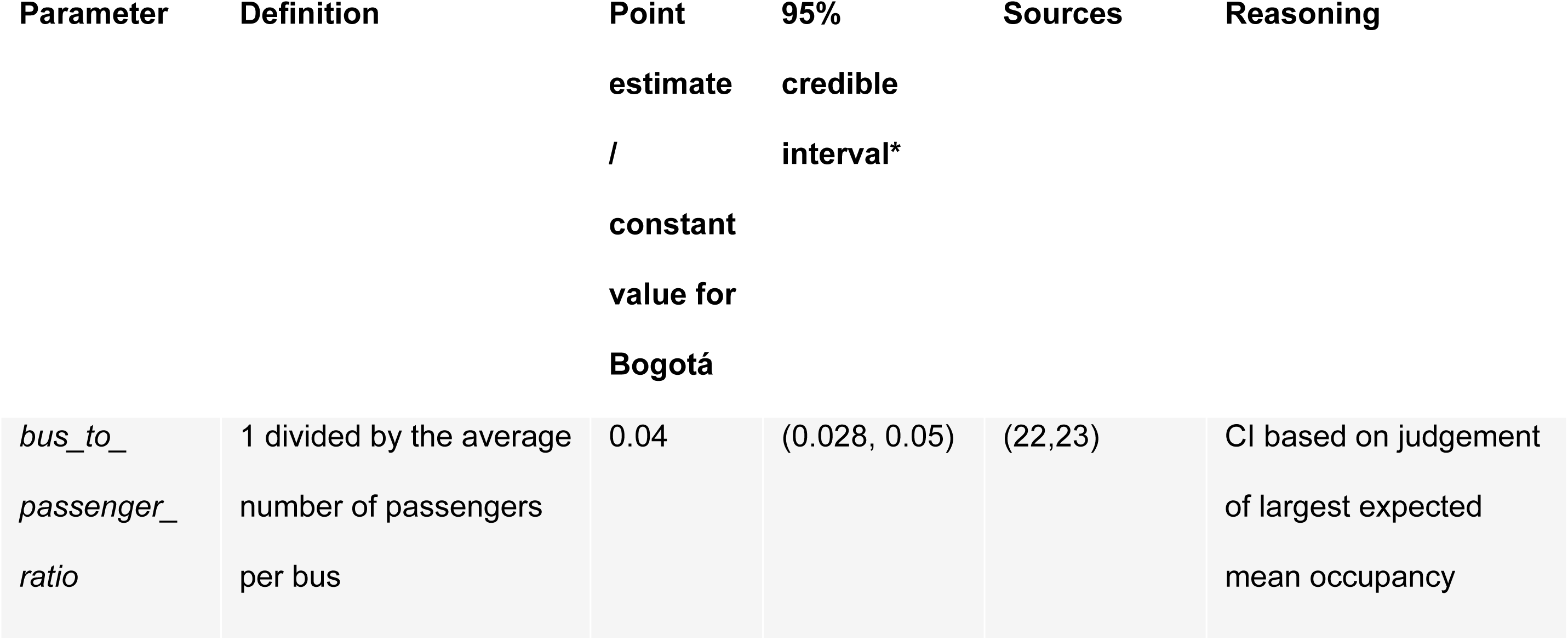
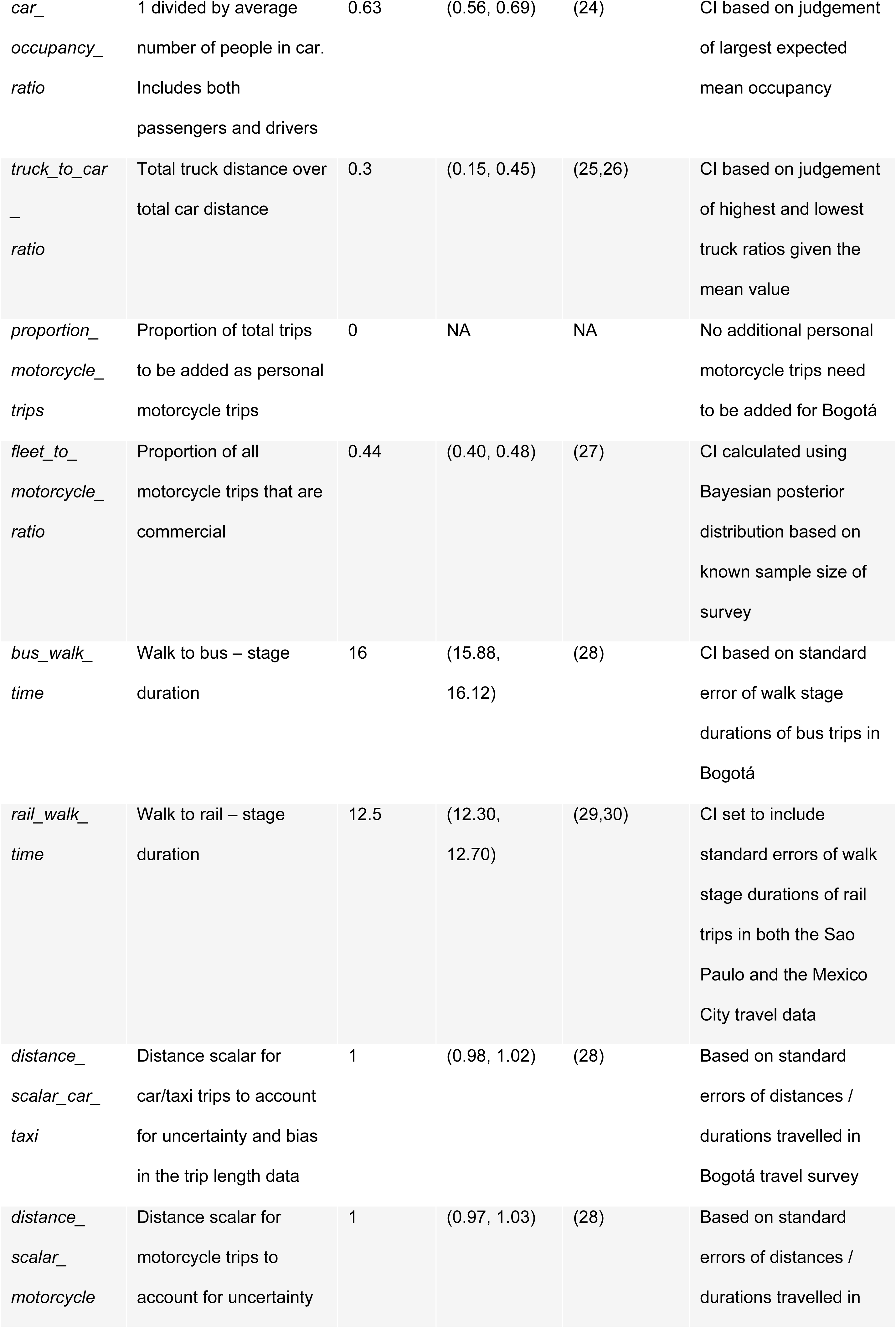
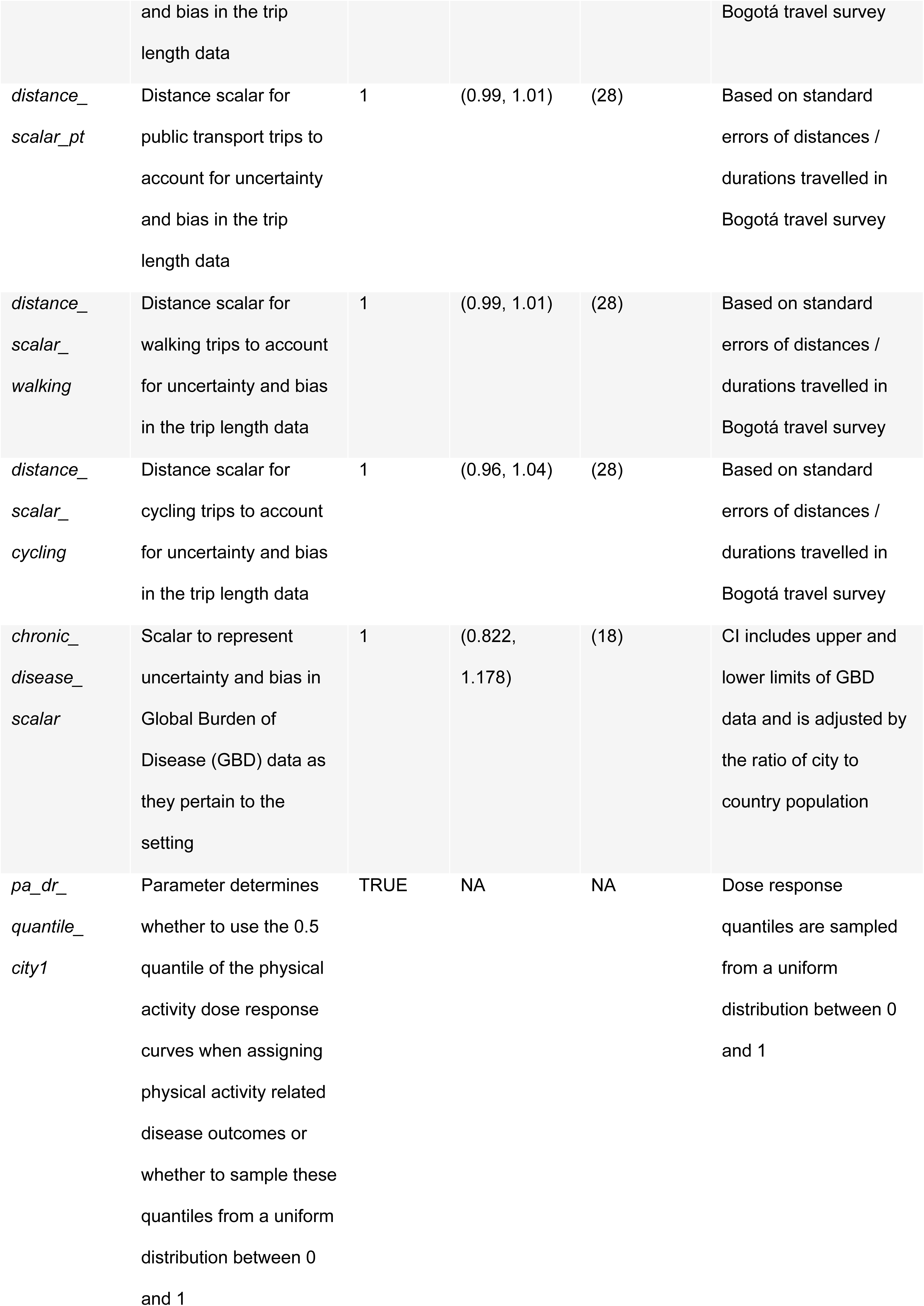
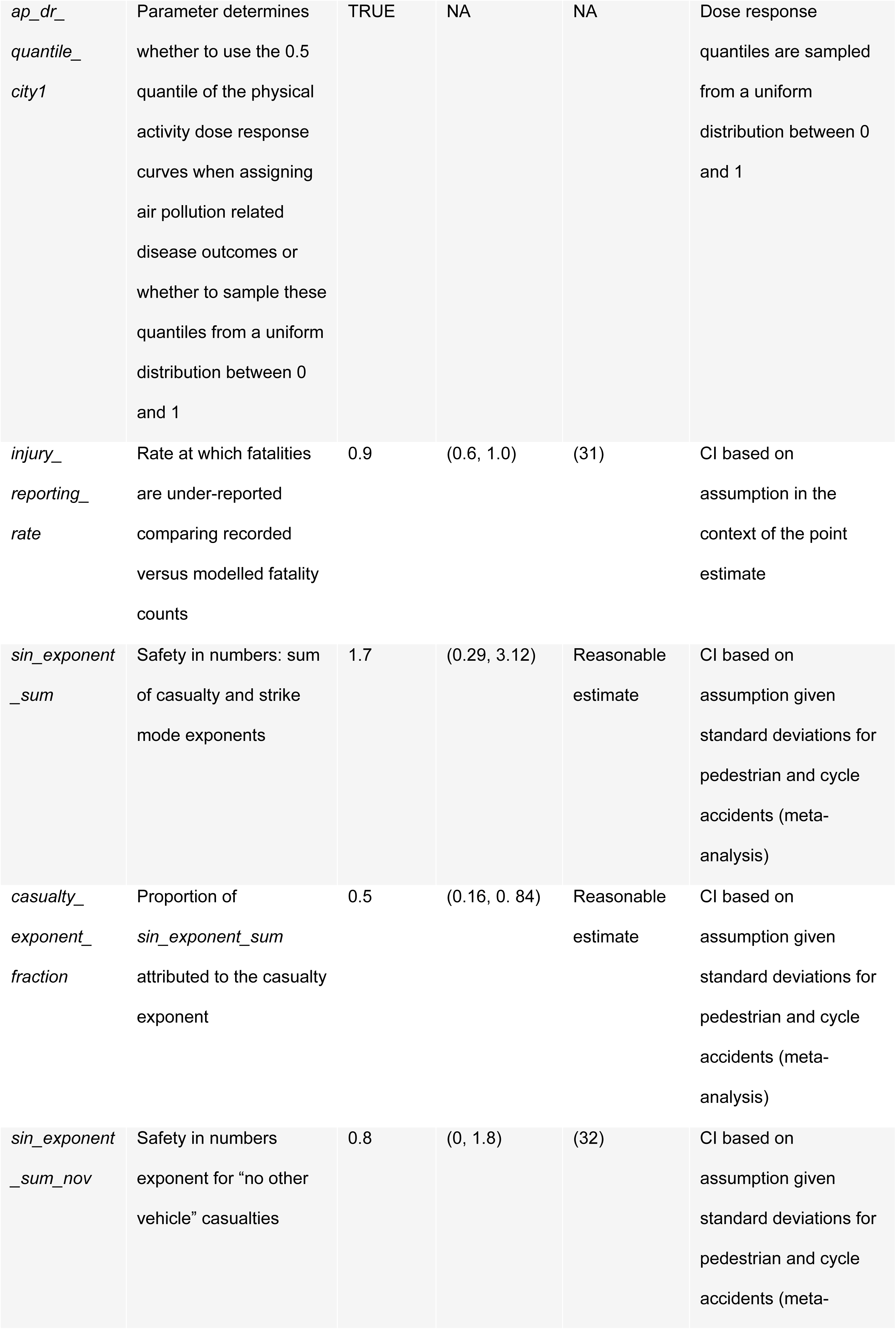
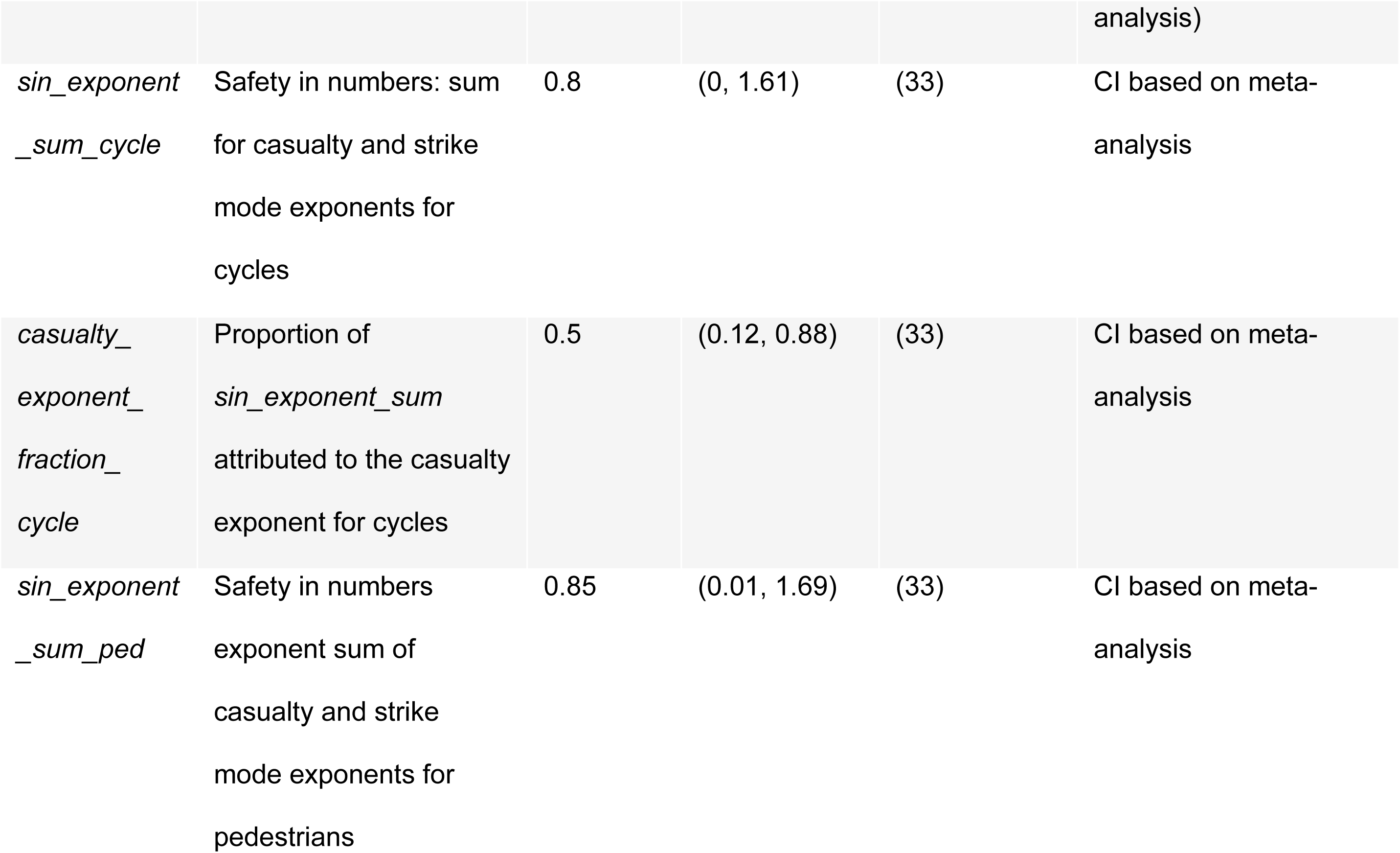
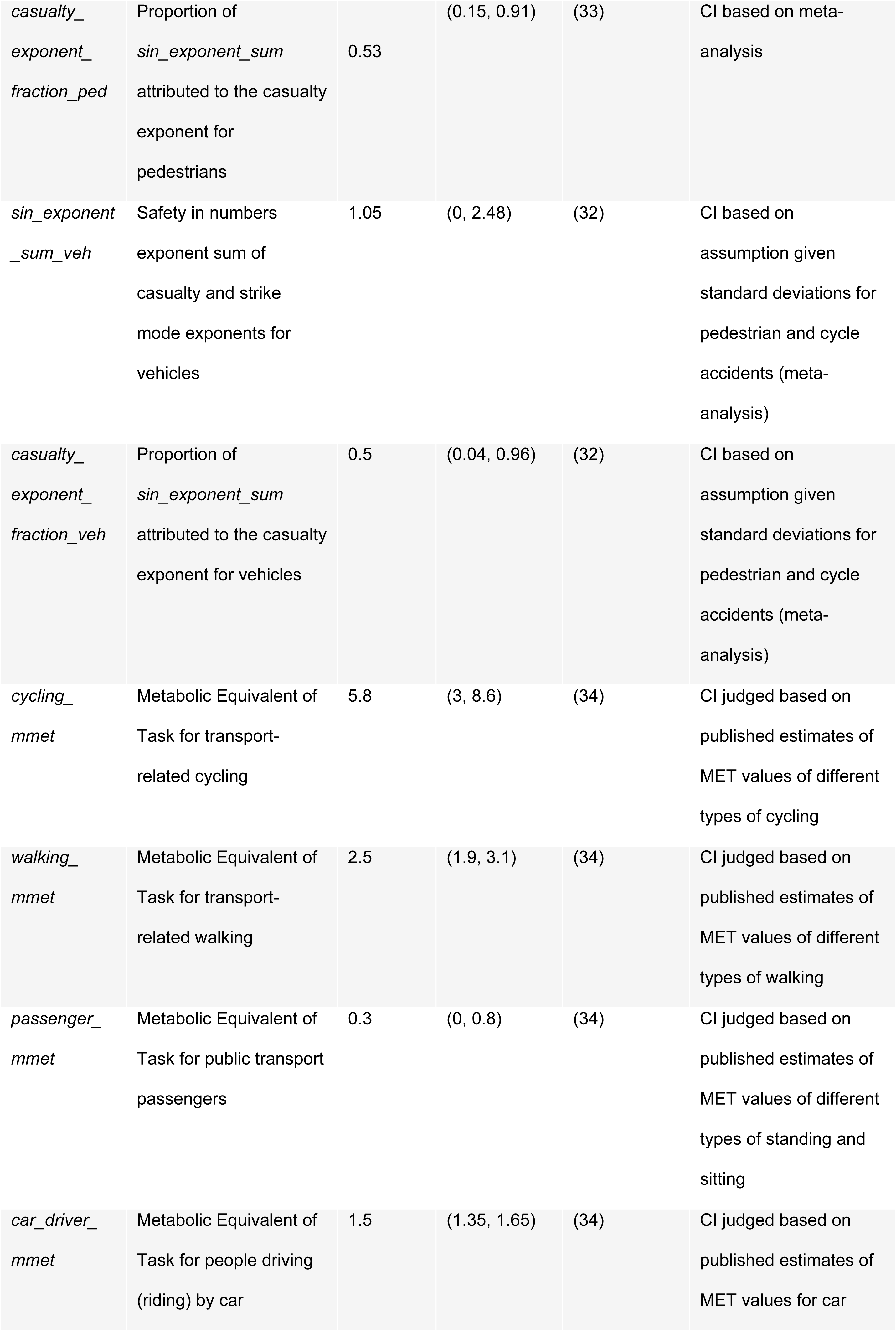
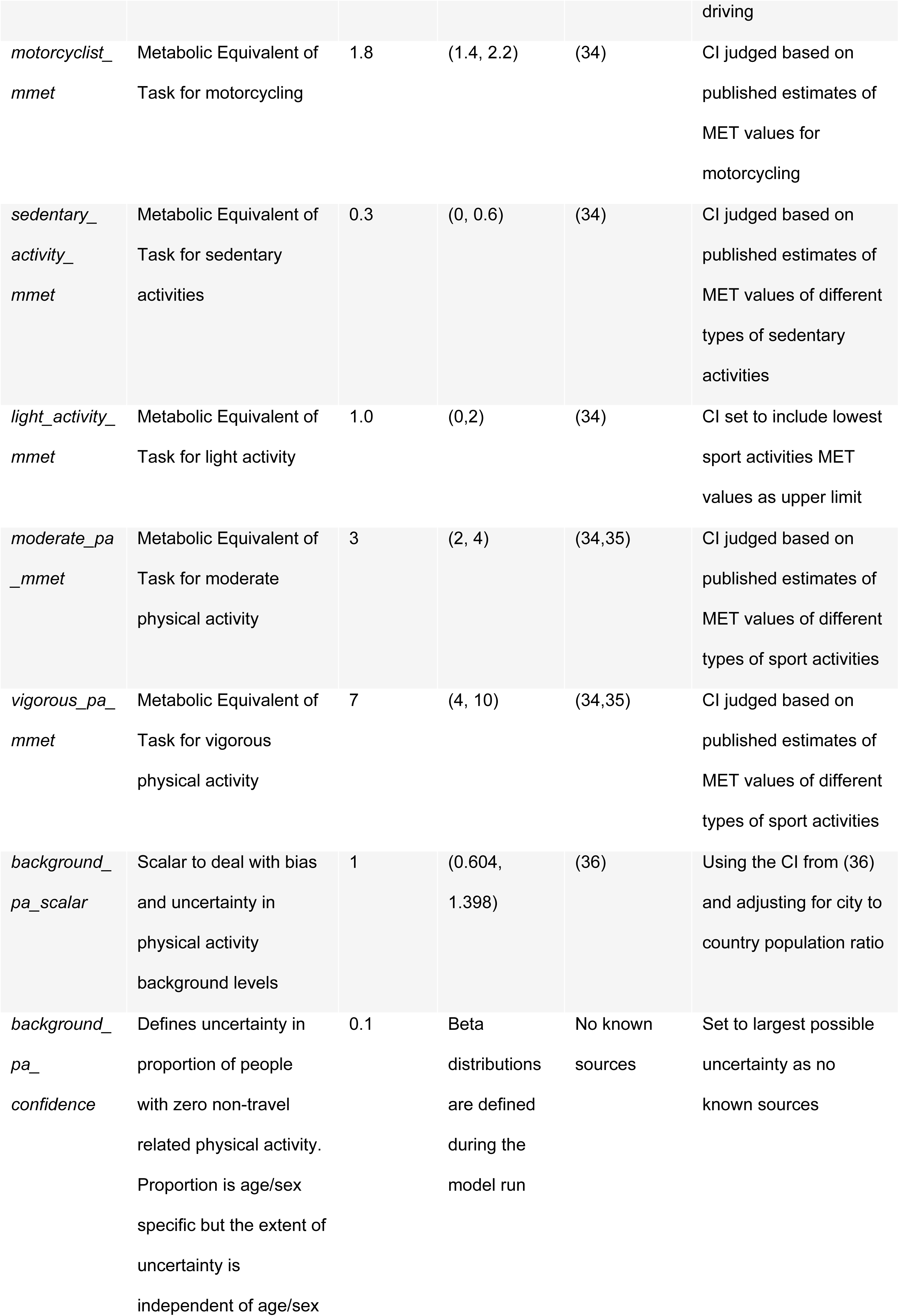
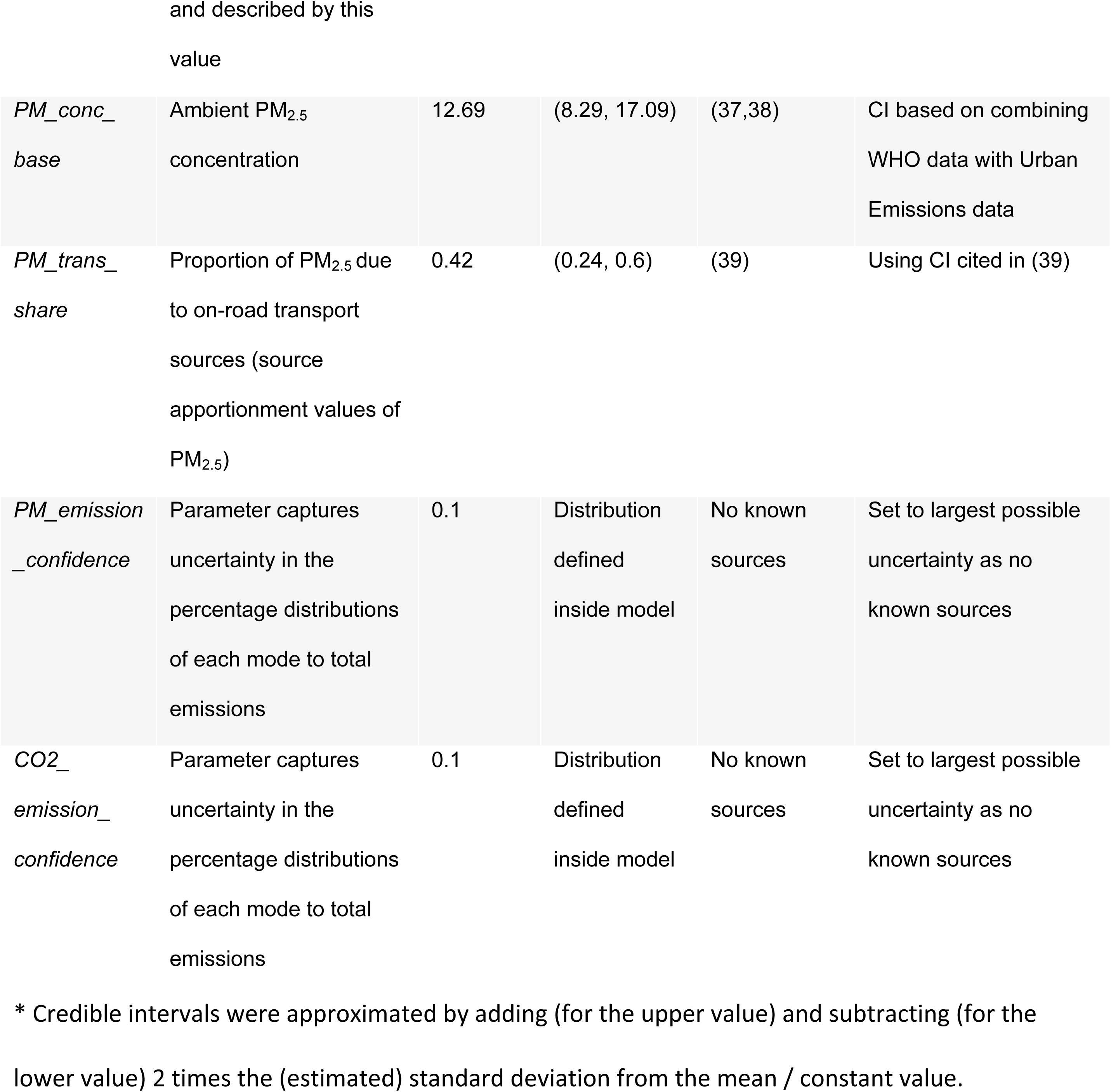
Input parameters with their credible intervals.

## 5. Results

We present two sets of results. The first set summarises the outputs from the Monte Carlo runs, i.e. YLL credible intervals for the health outcomes of interest. The second set describes the VoI results, which reflects the influence of the different input parameters on the uncertainty of the results.

### 5.1 Credible intervals from Monte Carlo simulations

The main output from the Monte Carlo simulation is the estimated change in YLLs for the different health outcomes of interest across the three scenarios. Fig 2 shows the change in total number of YLLs in level 1, i.e. air pollution and physical activity all-cause mortality and road traffic fatalities, for each scenario compared to the baseline. The bus scenario offers the highest benefits followed by the cycling scenario whereas the car scenario leads to a deterioration of health at the city level. The cycling scenario has the highest uncertainty with the 95% credible interval values lying between - 77% and +82% of the mean, whereas the bus scenario has the lowest levels of uncertainty with the 95% credible interval values lying between -41% and +52%. The width of the credible intervals demonstrates how uncertainty in the input parameters propagates through the model to the results. While uncertainty does not change whether a scenario is estimated to be harmful or beneficial (since each interval excludes zero), the estimated *size* of each effect spans a wide range of values.

**Fig 2.**
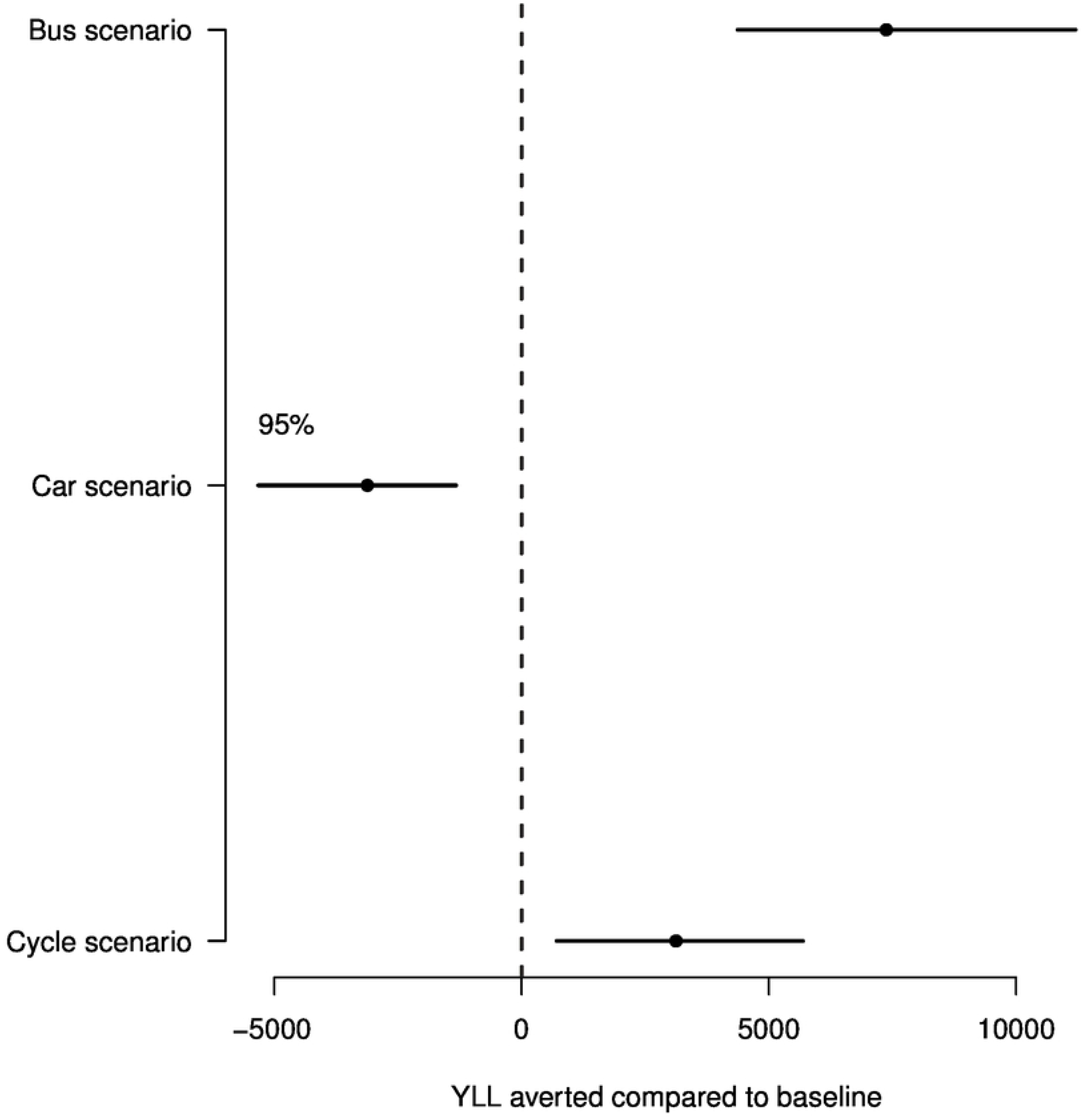
Changes in total YLL across the different scenarios for outcome level 1 (air pollution and physical activity all-cause mortality and road traffic fatalities) in Bogotá. Points show mean values. Lines show 95% credible intervals.

Disaggregated level 1 outputs (combined physical activity / air pollution (PA/AP) all-cause mortality and injuries results) demonstrate that the impacts of physical activity and air pollution on all-cause mortality exceed those of road-traffic injuries (Fig 3). Fig S2 shows that the physical activity / air pollution all-cause mortality outcomes are dominated by the physical activity contribution and air pollution does not have a large effect. Fig 3 also shows that impacts are greater for males across all scenarios, which occurs because the all-cause YLL burden in the baseline scenario is twice as large in the male population (18). One exception are road traffic fatality outcomes in the cycling scenario where females are more affected than male. The reason for this is that there are an additional 2470 female cycling trips compared with 1858 male cycling trips in the cycling scenario. These additional cycling trips result in 13 additional cycle fatalities (both sexes) but only 5 fewer pedestrian and 3 fewer motorcycle fatalities. As such there are more female fatalities in the cycling scenario.

**Fig 3.**
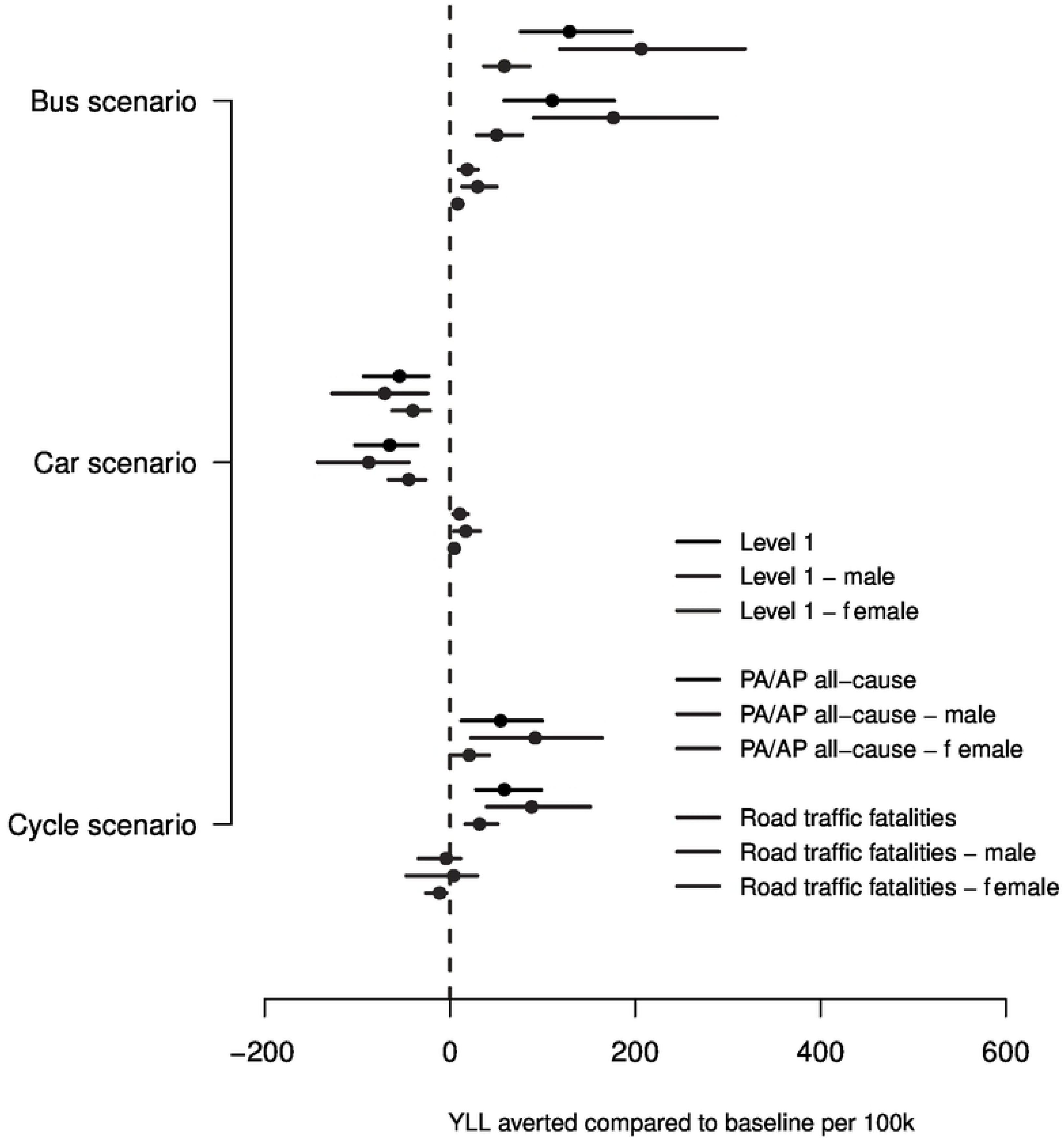
Changes in YLL per 100k people in Bogotá due to all-cause mortality and road traffic fatalities combined (level 1) and separately (combined air pollution (AP) and physical activity (PA) all-cause mortality and due road traffic fatalities). Each result is shown both aggregated and disaggregated by sex. Points show mean values. Lines show 95% credible intervals.

The following figure, Fig 4**E****rror! Reference source not found.**, shows the changes in YLLs per 100k people for all three outcome levels in total and by sex. Level 1 includes air pollution (AP) and physical activity (PA) all-cause mortality and road traffic fatalities, level 2 includes total cancer mortality (PA), cardiovascular diseases (AP and PA), respiratory diseases (AP) and road traffic fatalities, level 3 includes a variety of single disease outcomes for air pollution and physical activity and road traffic fatalities (see S8).

**Fig 4.**
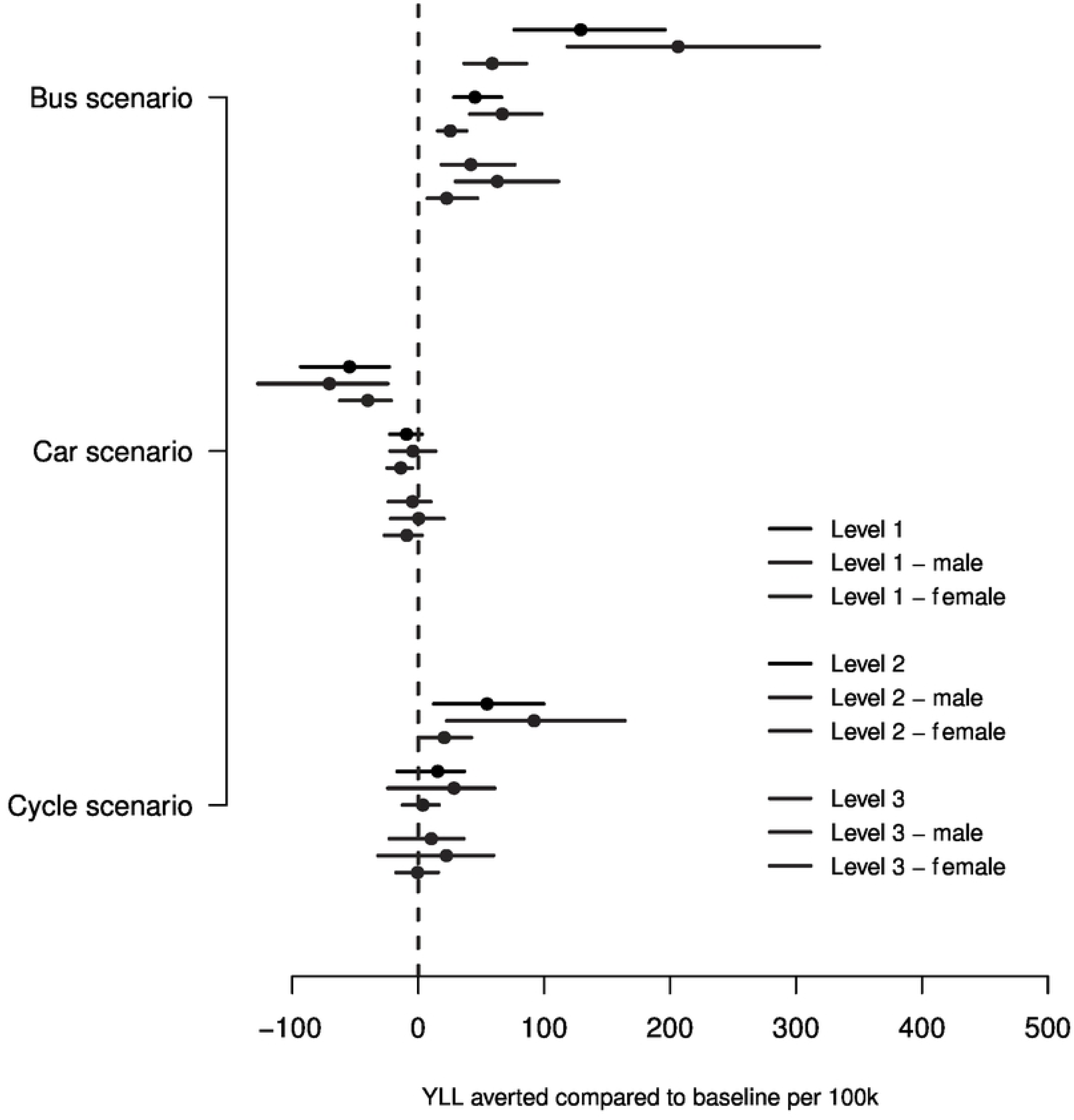
Changes in YLL per 100k for all three levels in total and disaggregated by sex in Bogotá. Level 1 includes combined air pollution (AP) and physical activity (PA) all-cause mortality and road traffic fatalities; level 2 includes combined disease outcomes for AP and PA such as cardiovascular diseases and road traffic fatalities and level 3 includes individual disease outcomes and road traffic fatalities. Points show mean values. Lines show 95% credible intervals.

As expected, we see the highest impacts at the highest level of abstraction (level 1: combined physical activity and air pollution all-cause mortality and road traffic fatalities). The estimated impact also has wider credible intervals than those at lower levels. Likely this is due to the uncertainty surrounding the physical activity and air pollution all-cause mortality outcomes (Fig 3), which has by far the largest standard deviation among all outcomes. The 95% credible intervals for the car and cycling scenarios at levels 2 and 3 include no change in YLL. Whilst the level 1 results are based on road traffic fatalities and the dose—response function results for all-cause mortality (one function), the level 2 and level 3 results are the sum of the outcomes of a variety of different diseases (and road traffic fatalities). These different disease outcomes can be either positively or negatively affected by changes in air pollution and physical activity levels. E.g. for level 2, all sexes, an increase in cycling has a mean negative effect on road traffic fatalities whereas it has a mean positive effect on total cancer rates. Adding these positive and negative outcomes to get the overall level 2 and level 3 results, it is not surprising that most of the resulting credible intervals in the car and cycling scenario contain 0 (a net neutral overall impact).

### 5.2 VoI analysis

In this section we look at the results from the VoI analysis to see which input parameters have the largest impacts on uncertainty in the output values. Fig 5 shows the 10 parameters which are expected to reduce the standard deviation for at least one of the outcomes physical activity/ air pollution all-cause mortality, road traffic fatalities, or both combined (level 1) for at least one of the three scenarios by at least 3% were we to know the true parameter value.

**Fig 5.**
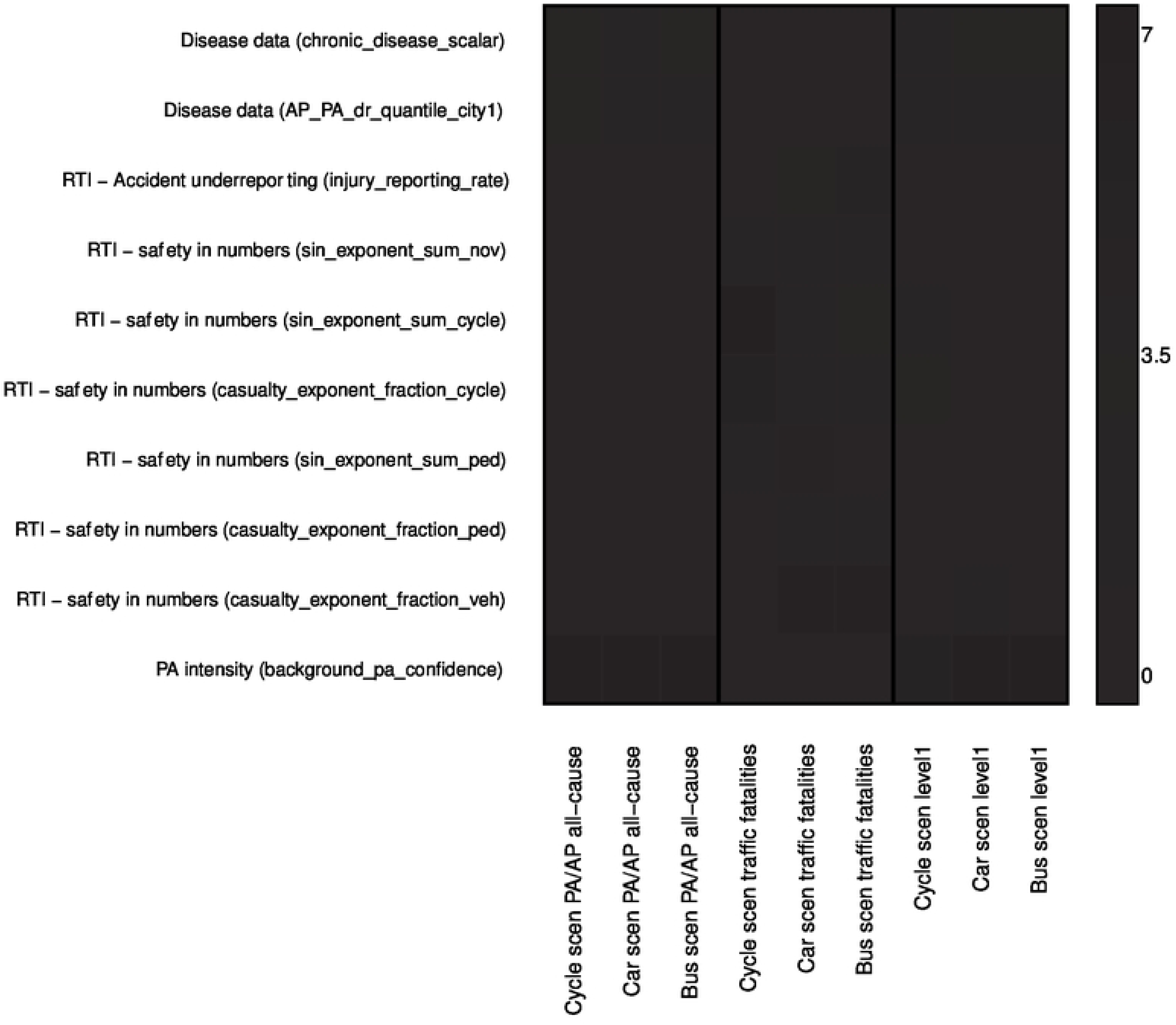
Parameters with the largest expected reduction in the standard deviation of air pollution (AP) and physical activity (PA) all-cause mortality, road traffic fatalities and level 1 results. Darker shading represents a higher percentage reduction in the standard deviation. RTI stands for road traffic injuries (fatalities only).

For the all-cause mortality and level 1 outcomes, better knowledge of some of the disease and physical activity data input parameters would have the largest effect. The most impactful disease data parameters consist of the “chronic disease scalar” which represents the unknown amount of bias and uncertainty associated with using the Global Burden of Disease data (S4.1). Knowledge of this parameter would reduce uncertainty due to air pollution and physical activity all-cause mortality in the car scenario by 3.6%. The disease parameter with the second largest effect is the *AP_PA_dr_quantile_city1* parameter which represents the uncertainty about the dose-response relationships (with perfect correlation assumed between all responses) (S4.2). The physical activity parameter with the largest effect is the “background PA confidence” which quantifies our uncertainty in the number of people with no non-travel related physical activity levels (S6.3). This parameter has by far the highest effect on air pollution and physical activity all-cause mortality and level 1 outcomes. Knowledge of this parameter would reduce outcome uncertainty for air pollution and physical activity all-cause mortality in the car scenario by up to 7%.

For the road traffic fatality results, the “safety in numbers” coefficients have the largest effect on uncertainty. These coefficients are used within the road traffic fatalities pathway to create a safety in numbers effect: the number of fatalities per distance for a mode goes down as the distance covered by that mode increases (S5.2). Additionally, the injury reporting rate, which quantifies our confidence in the observed accident fatality data (S5.1), has a substantial effect on uncertainty in road traffic fatalities for the car (3.4%) and bus (4.9%) scenarios.

Fig S3 shows the VoI analysis outcomes for all input parameters. The impact on the uncertainty of the results is negligible for most input parameters.

Fig 6 shows the six input parameters that, were we to know their true value, would be expected to reduce the uncertainty in physical activity and air pollution all-cause mortality, road traffic fatalities or both, for at least one of the three scenarios and at least one sex by at least 3%. Whilst all these parameters were also impactful for the aggregated results (Fig 5), not all of the parameters in Fig 5 have an effect of at least 3% when looking at the results split by sex. For the safety in number and injury reporting parameters there are substantial differences by sex, likely because the road traffic fatality rates by mode vary between males and females.

**Fig 6.**
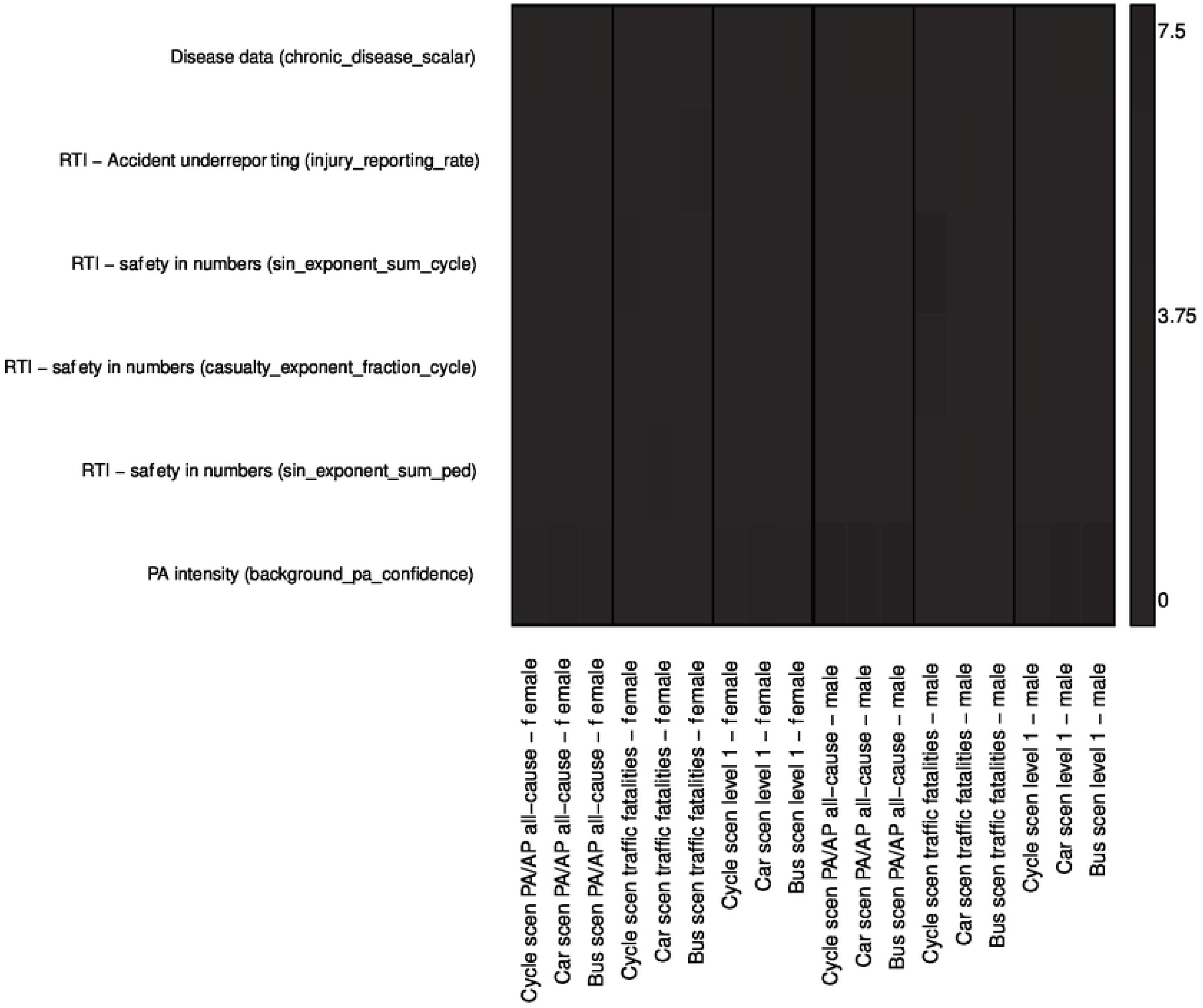
Parameters with the largest expected reduction in standard deviation of the air pollution (AP) and physical activity (PA) all-cause mortality, road traffic fatality and level 1 results by sex. Darker shading represents a higher percentage reduction in the standard deviation. RTI stands for road traffic injuries (fatalities only).

Fig S4 shows the effect of all input parameters on the results split by sex. Again, the impact of the majority of the parameters on our uncertainty about these health impacts is negligible.

## 6. Conclusion and further work

In this work, we present a case study of ITHM-Global in Bogotá: specifically, we show how it can be used for a substantive health impact analysis including uncertainty and Value of Information (VoI) analysis. We illustrate the use of ITHIM-Global in “sampling mode”, showing how to set up input parameter distributions, explaining the use of Monte Carlo simulation, and introducing VoI analysis. We present credible intervals for numerous modelled outcomes, and we identify parameters that most influence the widths of these credible intervals. We used Bogotá as our example city and modelled health impacts using three illustrative scenarios defined by a 5% increase in public transport, cycling, or car journeys. This work builds on a previous publication (13) that introduced ITHIM-Global, described its components, and how it can be used in “constant mode”, where all parameters are constant and uncertainties are excluded.

By running ITHIM-Global in sampling mode, we simultaneously take into account uncertainty in many input parameters. This helps us understand how uncertainty in the model outcomes is driven by uncertainty in all the considered input parameters at the same time, in contrast to one-way sensitivity analyses, where the uncertainty in the results is considered for one parameter at a time. VoI analysis allows us to determine the input parameters with the largest impact on output standard deviations, lending itself naturally to research prioritisation. We have presented this for overall results and by sex. The analysis could also be undertaken by age group and, where data are available, by socio-economic status.

In the Bogotá analysis, we find that the total uncertainty in the model output is large; for example, the reduction in YLLs in the bus scenario has a 95% credible interval of [4400 to 11,200] (Fig 2). Our VoI analysis estimates the reductions in this uncertainty that could be achieved if we knew the values of particular input parameters. While in reality we would not be able to obtain perfect information on any parameter, the VoI provides a useful upper bound for the potential gains in information. There is a large number of model parameters, each associated with a small proportion of the overall uncertainty (up to 7%), though the cumulative effect of learning many parameters would be higher. Note that despite the uncertainty in the *size* of the effects, for the main results (YLLs for [all-cause mortality and road traffic fatalities], Fig 2), we have shown that the *direction* of the effect is not affected by parameter uncertainty. We do not know, however, whether the bus scenario estimate is always bigger than the cycling scenario depending on the input parameters. Thus, the uncertainty analysis has improved our confidence that the bus and cycle scenarios are beneficial, and the car scenario is harmful. While in general our quantifications of input parameter uncertainty were based on judgements rather than precise data analyses, we aimed to err on the side of overestimating rather than underestimating uncertainty.

Further work should include more input parameter distributions, e.g. for speeds which are currently assumed to be constant. For those parameters with the largest expected impacts on key uncertainties, such as the proportion of people with no non-travel physical activity, the expected value of sample information might be used to predict the reduction in uncertainty that would be expected if we conducted a research study to estimate this parameter, such as a survey with a specific design and sample size. Whilst the VoI analysis is currently only set up to look at the impact on the joint results from the air pollution and physical activity pathways, it is of value to look at these two pathways separately as the results from the physical activity pathway currently dominate the results from the air pollution pathway.

Note that a VoI calculation refers to one specific model and output, describing the expected reductions in uncertainty if that model were used again to estimate the same output but with better information. However, better information on any parameter would have wider benefits beyond one model. Parameters in health impact models such as ITHIM can generally be classified as "local" or "global". In many cities and countries, there is a lack of local information on health-related exposures and outcomes, which can make modelling in that context more challenging. On the other hand, better information on global parameters, such as dose-response relationships, that are thought to be more consistent across all cities and countries, would improve health impact modelling in general.

Using a VoI analysis helps us to deal with the often high levels of uncertainty in data in low- and middle-income countries and as such helps to address inequities in health impact assessments between the Global North and the Global South.

## Data Availability

Population data comes from “Population projections disaggregated by locality 2018-2035” based on the National Population and Housing Census 2018 (https://www.dane.gov.co/index.php/estadisticas-por-tema/demografia-y-poblacion/proyecciones-de-poblacion/proyecciones-de-poblacion-bogota), travel behaviour comes from the 2019 Mobility survey from the Integrated Information System on Regional Urban Mobility (https://www.simur.gov.co/encuestas-de-movilidad), physical activity from the 2015 Colombian National Nutrition survey from the Ministry of Health (accessible upon request), road traffic injuries from the police department shared by the Bogotá Secretary of Mobility (https://datos.movilidadbogota.gov.co/search?groupIds=d3812f8315054cdc84cf744680103713), air pollution data from the 2022 version of WHO Air Quality Database (https://www.who.int/data/gho/data/themes/air-pollution/who-air-quality-database), vehicle emissions from EDGAR shared by Monica Crippa from the European Commission's Joint Research Centre, and baseline mortality and years of life lost data from the Global Burden of Disease study (GBD) published by the Institute for Health Metrics and Evaluation (https://ghdx.healthdata.org/gbd-2019).

https://www.who.int/data/gho/data/themes/air-pollution/who-air-quality-database

https://datos.movilidadbogota.gov.co/search?groupIds=d3812f8315054cdc84cf744680103713

https://www.simur.gov.co/encuestas-de-movilidad

https://www.dane.gov.co/index.php/estadisticas-por-tema/demografia-y-poblacion/proyecciones-de-poblacion/proyecciones-de-poblacion-bogota

## Acknowledgement

NA

## Supporting Information

**S1 Input parameters overview**

**S1 Table S1 Transport related input parameters**

**S1 Table S2 Health related input parameters**

**S1 Table S3 Road traffic fatalities related input parameters**

**S1 Table S4 Physical activity related input parameters**

**S1 Table S5 Air pollution related input parameters**

**S2 Choosing input parameter distributions**

**S2.1 Lognormal distributions**

**S2.2 Beta distributions**

**S2.3 Confidence parameters**

**S3 Transport related input parameters**

**S3.1 Bus to passenger ratio**

**S3.2 Car occupancy ratio**

**S3.3 Truck to car ratio**

**S3.4 Proportion motorcycle trips**

**S3.5 Fleet to motorcycle ratio**

**S3.6 Bus walk time and rail walk time**

**S3.7 Distance scalars**

**S3.7 Table S6 Distance scalar distributions**

**S3.8 Distributions of transport related input parameters**

**S3.8 Table S7 Parameters of transport related input parameter distributions**

**S4 Chronic burden of disease input parameters**

**S4.1 Chronic disease scalar**

**S4.2 Air pollution and physical activity dose response functions S4.3 Distributions of disease related input parameters**

**S4.3 Table S8 Parameters of disease related input parameter distributions**

**S5 Road traffic accident input parameters**

**S5.1 Injury reporting rate**

**S5.2 Safety in Number exponent sums and casualty exponent fractions for all modes**

**S5.2 Table S9 Constant casualty and strike mode exponents**

**S5.2 Table S10 Standard deviations of safety in number coefficients of walking and cycling accidents**

**S5.2 Table S11 Safety in number exponent sum input parameters**

**S5.2 Table S12 Safety in number casualty exponent fraction input parameters**

**S5.3 Distributions of road traffic accidents related input parameters**

**S5.3 Table S13 Parameters of road traffic accidents related input parameter distributions**

**S6 Physical activity input parameters**

**S6.1 MMET values**

**S6.1.1 Cycling**

**S6.1.2 Walking**

**S6.1.3 Passengers**

**S6.1.4 Car drivers**

**S6.1.5 Motorcycling**

**S6.1.6 Sedentary activity**

**S6.1.7 Light activity**

**S6.1.8 Moderate physical activity**

**S6.1.9 Vigorous physical activity**

**S6.2 Background physical activity scalar**

**S6.3 Background physical activity confidence**

**S6.3 Fig S1 Background physical activity confidence values.** Red shows the outputs for the various quantiles if the *background_pa_confidence* scalar is 0.1, green indicates a scalar of 0.5 and blue of 0.9. The doted lines give the 2.5^th^ quantiles, the solid lines the 50^th^ and the dashed lines the 97.5^th^ quantiles.

**S6.4 Distributions of physical activity related input parameters**

**S6.4 Table S14 Parameters of physical activity related input parameter distributions**

**S7 Air pollution input parameters**

**S7.1 PM_2.5_ concentration baseline**

**S7.2 PM_2.5_ transport share**

**S7.3 PM_2.5_ and CO_2_ emissions confidence**

**S7.4 Distributions of air pollution related input parameters**

**S7.4 Table S15 Parameters of air pollution related input parameter distributions**

**S8 Different disease levels considered in ITHIM-Global**

**S8 Table S16 Different disease levels considered in ITHIM-Global**

**S9 Running the VoI analysis script in ITHIM-Global**

**S9 Table S17 Input parameters defined in multi_city_voi.R script**

**S10 Results**

**S10.1 Further Monte Carlo simulations results**

**S10.1 Fig S2 Change in total YLL per 100k for level 1 (air pollution (AP) and physical activity (PA) all-cause mortality and road traffic fatalities), PA all-cause, AP all-cause mortality and road traffic fatalities.** Each result is shown both aggregated and disaggregated by sex. Points show mean values. Lines show 95% credible intervals.

**S10.2 Further EVPPI results**

**S10.2 Expected reduction in standard deviation of air pollution (AP) and physical activity (PA) all-cause mortality, road traffic fatalities and level 1 (PA/AP all-cause mortality and road traffic injuries) results in Bogotá were we to know the true value of the input parameters.** Darker shading represents a higher percentage reduction in the standard deviation. RTI stands for road traffic injuries (fatalities only).

**S10.2 Fig S 1. Expected reduction in standard deviation of air pollution (AP) and physical activity (PA) all-cause mortality, road traffic fatality and level 1 (PA/AP all-cause mortality and road traffic injuries) results in Bogotá by sex were we to know the true value of the input parameters.** Darker shading represents a higher percentage reduction in the standard deviation. RTI stands for road traffic injuries (fatalities only).

**S11 Choosing the Monte Carlo sample size**

**S11 Table S18 Uncertainty in results caused by different sample sizes**

**S11 Fig S5 Comparison of input parameters with largest EVPPI values for different sample sizes. AP stands for air pollution, PA for physical activity, RTI for road traffic injuries (fatalities only) and level 1 includes AP and PA all-cause mortality and road traffic fatalities.**

**S12 References**

**Fig S1.**
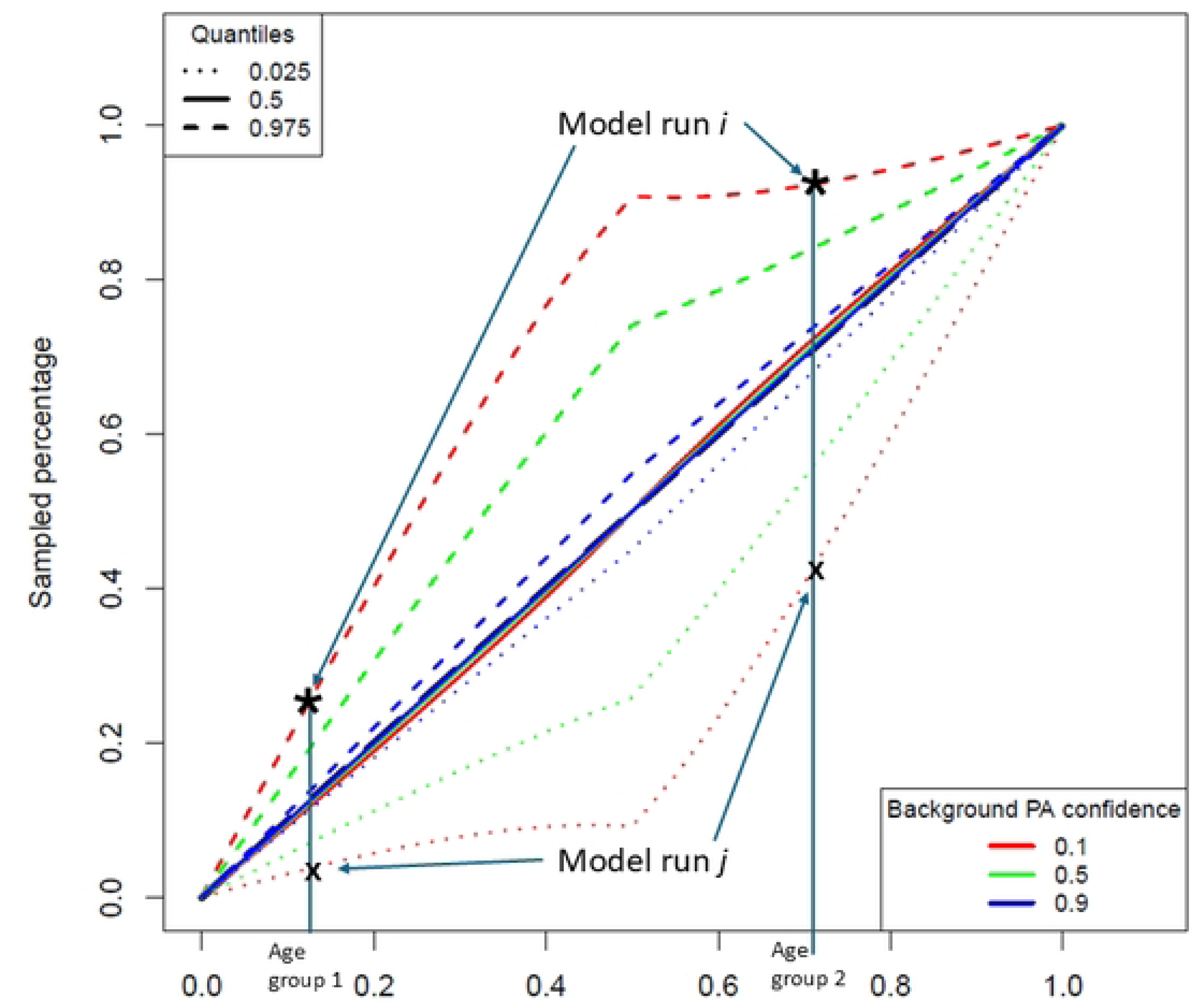

**Fig S2.**
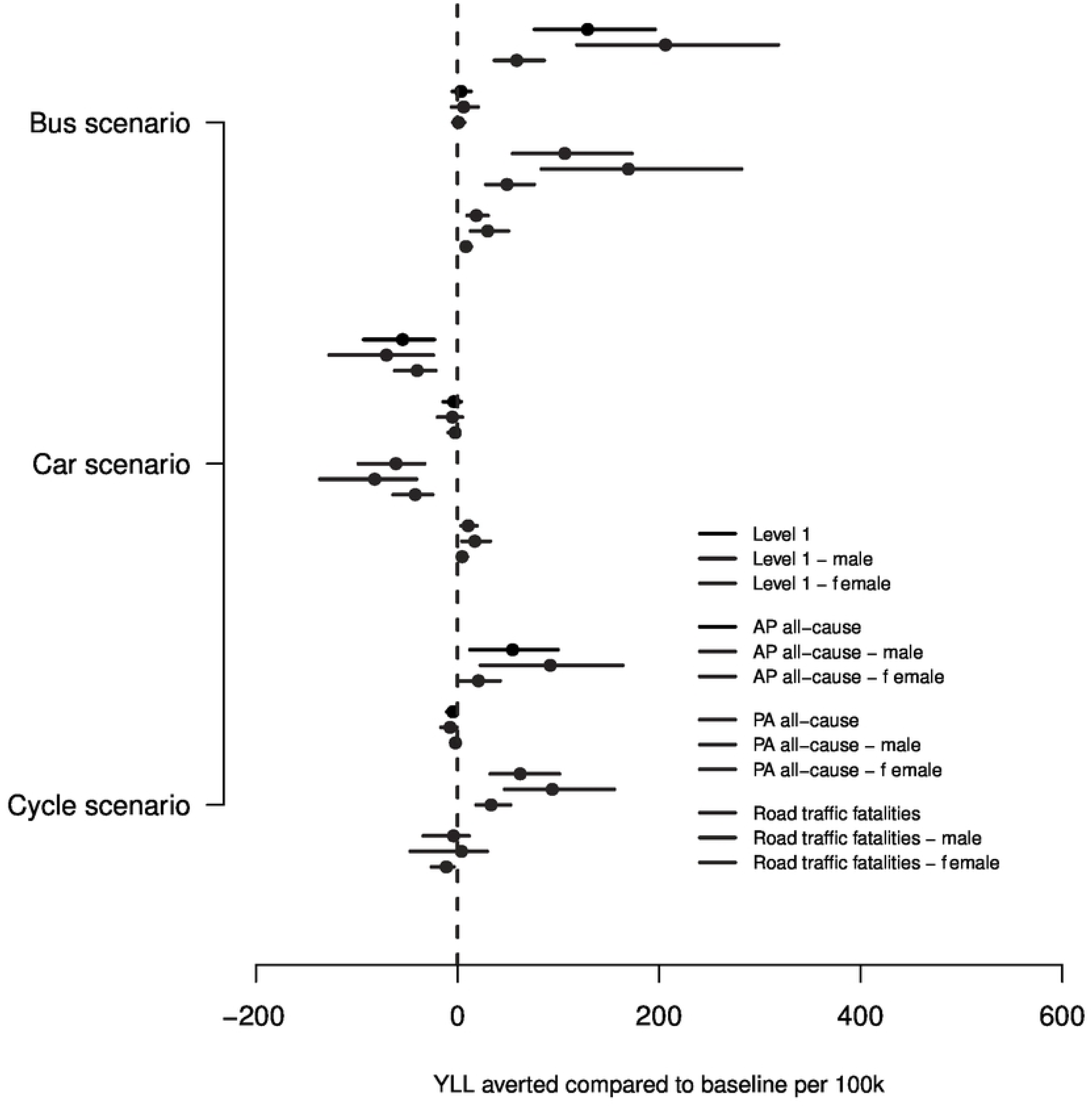

**Fig S3.**
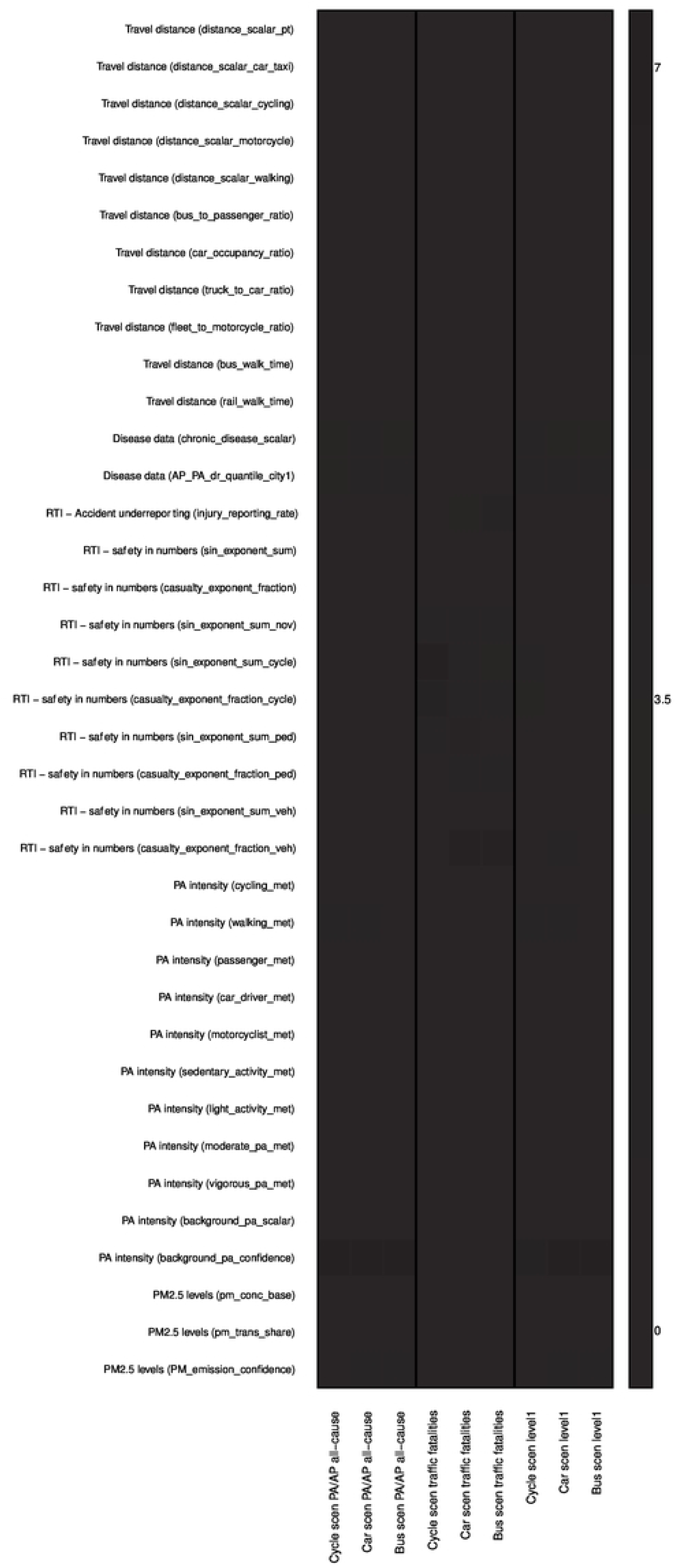

**Fig S4.**
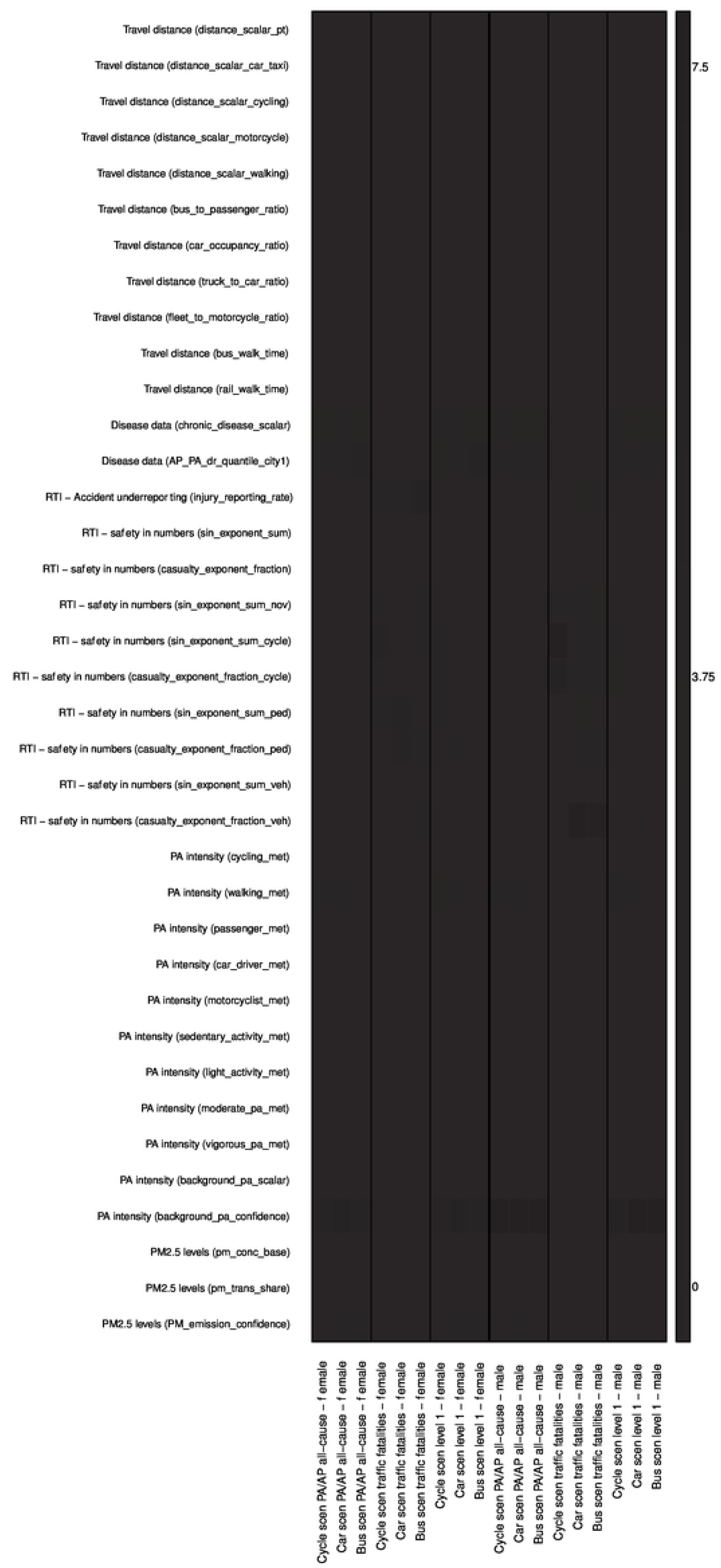

**Fig S5.**
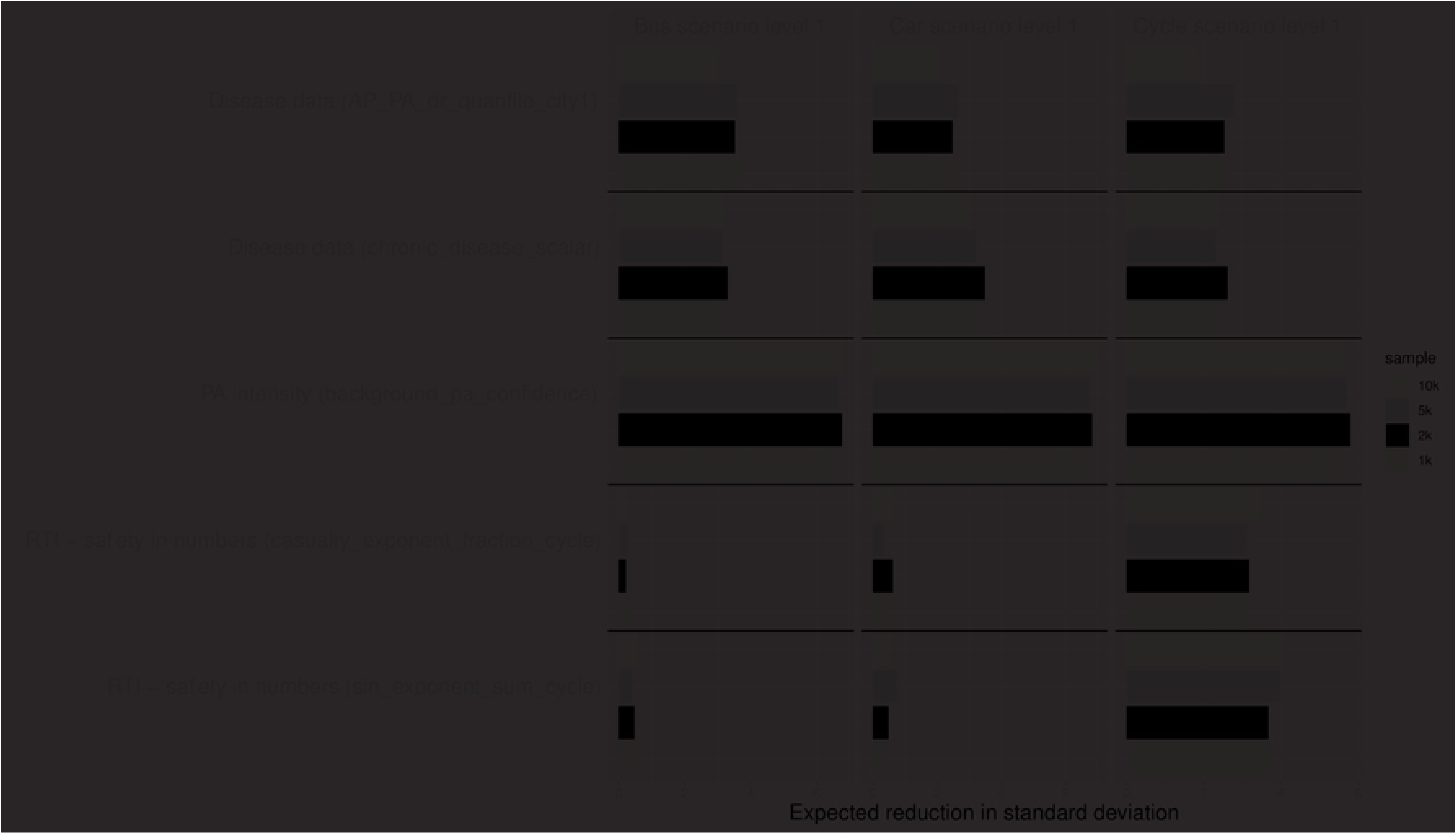

